# Survey data yields improved estimates of test-confirmed COVID-19 cases when rapid at-home tests were massively distributed in the United States

**DOI:** 10.1101/2024.05.21.24307697

**Authors:** Mauricio Santillana, Ata A. Uslu, Tamanna Urmi, Alexi Quintana, James N. Druckman, Katherine Ognyanova, Matthew Baum, Roy H. Perlis, David Lazer

## Abstract

**Importance:** Identifying and tracking new infections during an emerging pandemic is crucial to design and deploy interventions to protect populations and mitigate its effects, yet it remains a challenging task.

**Objective:** To characterize the ability of non-probability online surveys to longitudinally estimate the number of COVID-19 infections in the population both in the presence and absence of institutionalized testing.

**Design:** Internet-based non-probability surveys were conducted, using the PureSpectrum survey vendor, approximately every 6 weeks between April 2020 and January 2023. They collected information on COVID-19 infections with representative state-level quotas applied to balance age, gender, race and ethnicity, and geographic distribution. Data from this survey were compared to institutional case counts collected by Johns Hopkins University and wastewater surveillance data for SARS-CoV-2 from Biobot Analytics.

**Setting:** Population-based online non-probability survey conducted for a multi-university consortium —the Covid States Project.

**Participants:** Residents of age 18+ across 50 US states and the District of Columbia in the US.

**Main Outcomes and Measures:** The main outcomes are: (a) survey-weighted estimates of new monthly confirmed COVID-19 cases in the US from January 2020 to January 2023, and (b) estimates of uncounted test-confirmed cases, from February 1, 2022, to January 1, 2023. These are compared to institutionally reported COVID-19 infections and wastewater viral concentrations.

**Results:** The survey spanned 17 waves deployed from June 2020 to January 2023, with a total of 408,515 responses from 306,799 respondents with mean age 42.8 (STD 13) years; 202,416 (66%) identified as women, and 104,383 (34%) as men. A total of 16,715 (5.4%) identified as Asian, 33,234 (10.8%) as Black, 24,938 (8.1%) as Hispanic, 219,448 (71.5%) as White, and 12,464 (4.1%) as another race. Overall, 64,946 respondents (15.9%) self-reported a test-confirmed COVID-19 infection. National survey-weighted test-confirmed COVID-19 estimates were strongly correlated with institutionally reported COVID-19 infections (Pearson correlation of r=0.96; p=1.8 e-12) from April 2020 to January 2022 (50-state correlation average of r=0.88, SD = 0.073). This was before the government-led mass distribution of at-home rapid tests. Following January 2022, correlation was diminished and no longer statistically significant (r=0.55, p=0.08; 50-state correlation average of r=0.48, SD = 0.227). In contrast, survey COVID-19 estimates correlated highly with SARS-CoV-2 viral concentrations in wastewater both before (r=0.92; p=2.2e-09) and after (r=0.89; p=2.3e-04) January 2022. Institutionally reported COVID-19 cases correlated (r = 0.79, p=1.10e-05) with wastewater viral concentrations before January 2022, but poorly (r = 0.31, p=0.35) after, suggesting both survey and wastewater estimates may have better captured test-confirmed COVID-19 infections after January 2022. Consistent correlation patterns were observed at the state-level. Based on national-level survey estimates, approximately 54 million COVID-19 cases were unaccounted for in official records between January 2022 and January 2023.

**Conclusions and Relevance:** Non-probability survey data can be used to estimate the temporal evolution of test-confirmed infections during an emerging disease outbreak. Self-reporting tools may enable government and healthcare officials to implement accessible and affordable at-home testing for efficient infection monitoring in the future.

**Trial Registration:** NA

**Key Points:** *Question:* Can non-probability survey data accurately track institutionally confirmed COVID-19 cases in the United States, and provide estimates of unaccounted infections when rapid at-home tests are popularized and institutionalized tests are discontinued?

*Findings:* The proportion of individuals reporting a positive COVID-19 infection in a longitudinal non-probability survey closely tracked the institutionally reported proportions in the US, and nationally-aggregated wastewater SARS-CoV-2 viral concentrations, from April 2020 to February 2022. Survey estimates suggest that a high number of confirmed infections may have been unaccounted for in official records starting in February 2022, when large-scale distribution of rapid at-home tests occurred. This is further confirmed by viral concentrations in wastewater.

*Meaning:* Non-probability online surveys can serve as an effective complementary method to monitor infections during an emerging pandemic. They provide an alternative for estimating infections in the absence of institutional testing when at-home tests are widely available. Longitudinal surveys have the potential to guide real-time decision-making in future public health crises.

## Introduction

Identifying and tracking new infections during the earlier and most intense phases of the COVID-19 pandemic was crucial for the design of mitigation strategies. Yet it was extremely challenging due to the novel nature of the pathogen ^1,2^. The significant number of asymptomatic COVID-19 infections, the limited availability of resources to identify and treat infections across locations, and people’s lack of trust and willingness to seek medical attention were some of the most important challenges of estimating incidence numbers ^3–6^. Multiple approaches to characterize the incidence of COVID-19 in the population were deployed in the US as infections spread ^7^ These included (a) clinical-based individual testing (via polymerase chain reaction, PCR, or rapid tests) ^8^, (b) tracking the number of patients in hospital visits with COVID-19 symptoms, such as fever, cough, sore throat, anosmia (referred to as “syndromic surveillance”)^9^; (c) the continuous monitoring of the presence of antibodies against SARS-CoV-2, the virus that causes COVID-19 infections, in the blood serum of a population (referred to as antigen testing and serosurveillance) ^3,10^ and (d) measuring the amount of SARS-CoV-2 viral concentration in wastewater samples shed by infected individuals ^4,11,12^.

Among all these, widespread institutional individual testing was the most heavily relied-upon indicator to determine the severity of local outbreaks, allocate resources, and deploy or lift non-pharmaceutical mitigation interventions. Throughout the pandemic, however, testing availability and reporting were inconsistent in the US ^13^. For example, the COVID-19 tests —designed by the US Centers for Disease Control and Prevention, (CDC)— were recalled due to a faulty reagent ^14^ during the earlier months of 2020, heterogenous state policies regarding access to free institutional testing led to inconsistencies in interpreting case count data^15^, and the massive government-led distribution of rapid at-home tests starting in January 2022, without a concurrent deployment of a centralized infection reporting system, meant low coverage took place.

Here, we study the ability of data collected from large US-based nonprobability surveys –the COVID States Project (CSP)– to estimate the number of COVID-19 infections from January 2020 to January 2023, at national and state levels. Multiple studies have investigated how surveys can be leveraged to monitor infections, people’s behaviors, and trust in vaccine in specific periods and particular geographies during the COVID-19 pandemic ^16,17^. In this study, we further sought to assess the extent to which carefully-analyzed survey data could have been used to monitor the number of COVID-19 infections *continuously* and *longitudinally* at the national and state levels during the first 3 years of the pandemic in the US.

## Methods

### Study Design

We used data collected by an ongoing large-scale internet-based nonprobability survey conducted by an academic consortium approximately every 6 weeks from April 2020 onwards, inclusive of all 50 states and the District of Columbia. Survey participants were individuals aged 18 years or older who resided in the United States. Importantly, before accepting to participate in the survey, respondents were not aware that the survey would include questions related to the COVID-19 pandemic, in order to minimize selection bias. The survey used national and state-level representative quotas for gender, age, and race/ethnicity to represent the US population in the most recent census data. Participants were recruited using PureSpectrum, an online survey panel aggregator, and they provided informed consent online before survey access.

From the 5th survey wave (June 2020) onwards, the surveys asked two questions to identify the COVID-19 test frequency of participants, positive test results, and the month when they experienced symptoms. The precise wording of the questions can be found in Supplemental Materials.

### Measures

All respondents were asked if they had been tested for COVID-19 in the past (not distinguishing between PCR test or antigen test in some waves), and those who indicated a positive test result were asked when they experienced symptoms. To estimate the number of infections happening in each month, we aggregated the number of respondents who indicated having a positive test result and were sick in each month, only using the immediately subsequent survey wave after each individual’s infection to minimize potential participants’ recall errors. About 16% of respondents participated in multiple survey waves, and if they reported multiple infections in different months, we included their health status in each month they reported an infection. Sensitivity analyses were conducted to test whether including more than one infection per respondent would yield different results compared to only including at most one (randomly selected) infection per respondent. The aggregated responses were demographically reweighted to represent the most recent US Census and normalized by the sample size, to estimate the proportion of infected individuals at the national and state levels. The sample sizes and percent of respondents who were sick in each month at the national level are shown in the Supplementary Materials. Institutionally-confirmed COVID-19 infections were obtained from state and local governments and health departments by the Coronavirus Resource Center of Johns Hopkins University (JHU), and compiled by the New York Times (<u>covid-19-data</u>). Finally, as an additional and independent measure of COVID-19 prevalence in the US, we used monthly aggregated wastewater SARS-CoV-2 viral concentrations from Biobot Analytics.^18^

### Statistical Analysis

We conducted our statistical analyses in two different and non-overlapping time periods within the first three years of the pandemic in the US, selected a priori. The first one was from April 1, 2020 to January 31, 2022, a time when institutional efforts to test individuals were most active –according to the number of daily PCR COVID-19 tests conducted --see Supplementary Materials. The second one, from February 1, 2022, to January 1, 2023, a time when rapid at-home tests were massively distributed to the general public by the federal government. During this period, there was not a centralized system to record the rapid test outcomes, and an overall decrease in governmental resources allocated to monitor COVID-19 infections gradually occurred; culminating with the federal Public Health Emergency for COVID-19 expiring in May 2023.

#### Correlation analysis

We calculated pairwise correlation coefficients between the proportion of infected individuals as inferred by survey data (referred to as CSP) and the institutional numbers reported in the Johns Hopkins University (JHU) COVID-19 dashboard, for the two time periods described above at the national and state levels. We also calculated pairwise correlations between SARS-CoV-2 viral load in wastewater and both the CSP estimates JHU reported infections, during the two distinct periods. This was done at the national and state levels.

#### Survey Mean Estimates and Standard Errors

We measured the distance between the official numbers and our survey estimates as multiples of the survey-based standard errors (standard deviation of mean) of our estimates. We report both these standardized differences and the 95% confidence intervals in Figure 1 for the national level.

**Figure 1.**
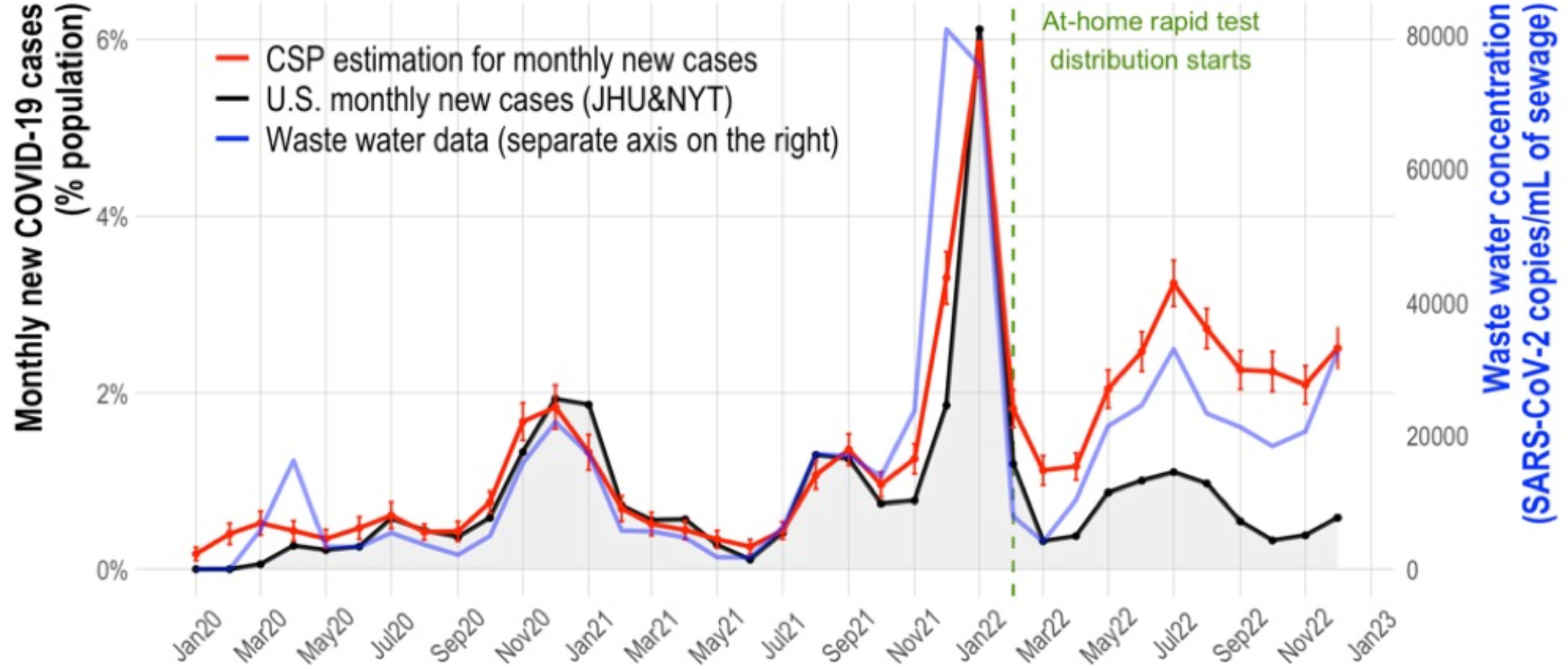
The percent of respondents in our survey who reported having a confirmed COVID-19 infection in each month is shown in red (CSP), the institutionally reported percent of individuals infected in each month as monitored by JHU is shown in black, and the wastewater viral concentration of SARS-CoV-2 is shown in blue. A vertical green dashed line shows the time when at-home rapid test were widely delivered in February 2022.

#### Excess infection estimates

We estimated the number of infections that were likely unaccounted for by institutional surveillance systems starting in February 2022 using two approaches. The first one involved calculating the cumulative infections estimated from survey data from February 1, 2022, until January 1, 2023. This was achieved by multiplying the percent of weight-corrected self-reported infected participants in each month by the total population of the US, and then by adding all these estimates across time. We then simply subtracted the cumulative number of infections reported by the JHU from the survey’s cumulative estimates. A second approach involved calibrating a linear regression model to map CSP COVID-19 estimated incidence onto JHU’s reported COVID-19 incidence from April 2020 to January 2022, to identify the relationship between the two in the first 2 years when they closely tracked each other. We then used this model to predict confirmed infections after February 2022 –with the assumption that we only had access to CSP information. We computed cumulative values for the predicted JHU that would have been observed in the absence of any disruption (intervention) to the institutional COVID-19 surveillance, and then compared these estimates to the cumulative cases calculated from JHU data. This method is detailed in De Salazar et al 2021 in the context of COVID-19 vaccine effectiveness, and is frequently referred to as interrupted time-series analysis.^19,20^

## Results

The survey spanned 17 waves deployed from June 2020 to January 2023, with a total of 408,515 responses from 306,799 respondents with mean age 42.8 (STD 13) years; 202,416 (66%) identified as women, and 104,383 (34%) as men. A total of 16,715 (5.4%) identified as Asian, 33,234 (10.8%) as Black, 24,938 (8.1%) as Hispanic, 219,448 (71.5%) as White, and 12,464 (4.1%) as another race. Overall, 64,946 respondents (15.9%) self-reported a test-confirmed COVID-19 infection. In aggregate and at the national level, COVID-19 case counts inferred from CSP surveys are highly correlated with JHU reports from April 2020 to January 2022 (Pearson r=0.96; p=1.8e-12), as seen in Figure 1 and Table 1 –with state-level Pearson correlations average of r=0.88 (STD 0.074) and all significant. After February 2022, soon after at-home rapid tests were massively distributed by the federal government, and up to January 2023, the Pearson correlation between CSP and JHU case counts dropped to r=0.55 (p=0.08) with state-level Pearson correlations average of r=0.48 (SD = 0.227) and mostly not statistically significant. Sensitivity analysis shown in the supplementary materials led to very similar temporal infection curves when including repeat participants only (randomly) once in our study.

**Table 1.** National-level pairwise Pearson correlation and p-values between survey test-confirmed infections estimates (CSP), Institutionally reported COVID-19 (JHU), and Wastewater SARS-CoV-2 viral concentrations (WW) in three time periods.

|  | Apr 2020 – Jan 2022<br>(pre-rapid test period) | Feb 2022 – Jan 2023<br>(rapid test period) | Apr 2020 – Jan 2023<br>(full time period) |
| --- | --- | --- | --- |
| **CSP – JHU** | 0.96 (p=1.8e-12) | 0.55 (p=0.08) | 0.78 (p=1.1e-07) |
| **CSP – WW** | 0.92 (p=2.2e-09) | 0.89 (p=2.33e-04) | 0.87 (p=3.8e-11) |
| **JHU – WW** | 0.79 (p=1.1e-05) | 0.31 (p=0.35) | 0.74 (p=1.1e-06) |

Additionally, Table 1 shows that concentrations of SARS-CoV-2 in wastewater data (WW) closely correlate with CSP surveys’ case counts both between April 2020 and January 2022 (r=0.92 p=2.2e-09), and between February 2022 and January 2023 (r=0.89 p=2.3e-04). While SARS-CoV-2 in wastewater (WW) correlates (r=0.79 p=1.10e-05) with JHU case counts before January 2022, this correlation drops dramatically to r= 0.31 (p=0.35) between February 2022 and January 2023. Consistent correlation patterns are observed at the state-level and are reported in the Supplementary Materials.

Using the first approach to calculate cumulative infections from February 1, 2022, to January 1, 2023 (post at-home tests distribution) at the national level, our survey estimates suggest that about 79 million (95% CI: 71 million – 86 million) confirmed cases may have occurred; compared to 25 million reported in the JHU data. This indicates that 54 million cases, more than twice as many of those reported, were likely unaccounted for in institutional surveillance. At the state level, the number of potentially unaccounted cases vary between 59K in Wyoming to 6.3 million in California. Our second (interrupted time-series) approach, the linear regression (JHU = 0.96*CSP-0.1) calibrated during the April 2020 to January 2022 nationally, yields consistent results, and predicts that the cumulative number of positive cases from February 1, 2022, to January 1, 2023 was 73 million (95% CI: 65 million – 81 million) at the national level. State-level results are consistent and shown in the Supplementary Materials.

For 22 months, from April 2020 when reliable institutionalized testing in the US accelerated, until February 2022, the official data fell within 2 to 3 SEs away from our estimates, except the months January, November, and December 2021. However, from February 2022 onward, when the distribution of rapid at-home tests started, the distance between survey estimates and the officially reported cases started to diverge significantly, ranging from 6 SEs in February 2022 to 16 SEs during the peak of July 2022.

## Discussion

Our results support the hypothesis that nonprobability surveys serve as a reliable and complementary method to monitor the proportion of test-confirmed infections in real-time during a public health crisis. Specifically, by analyzing data from nonprobability surveys deployed approximately every 6 weeks during the first three years of the pandemic in the US, we found that COVID-19 infections inferred from survey data closely tracked institutionally reported infections when institutional testing was at its best in the US. When institutional efforts to monitor COVID-19 infections diminished and rapid at-home tests were made widely accessible – with no centralized system to collect at-home test results – survey data suggested that a high proportion of test-confirmed infections were unaccounted for in institutional reports. When comparing with COVID-19 activity estimates obtained from SARS-CoV-2 concentrations in wastewater data, we find high consistency with the COVID-19 trends observed in surveys throughout this study.

The alignment of the three – JHU, CSP, and WW– COVID-19 activity estimates before January 2022 suggests that these surveillance systems were consistent and compatible with one another before the mass distribution of at-home tests. After February 2022, the consistency between CSP and WW COVID-19 activity, and the pronounced discordance between these two sources and JHU cases, suggest that both (a) CSP and WW data may have continued properly capturing COVID-19 infections trends, and (b) the introduction of at-home rapid tests and the discontinuation of institutional testing disrupted institutional efforts (JHU) to track COVID-19 trends. Similar alignment between the three data sources is observed before January 2022, at the state level. There also were clear discrepancies between both CSP and WW with JHU data after February 2022 at the state level, as shown in the Supplementary Materials.

While there have been multiple attempts to monitor or estimate the number of confirmed COVID-19 cases using alternative Internet-based data sources, such as digital Internet traces (e.g. general population’s Internet search queries, Clinicians’ searches, among others ^21^), human mobility data from smartphones ^22–24^, self-test reporting systems ^25^, and surveys like ours ^26^, our study presents among the most comprehensive assessments of the quality of COVID-19 activity estimates using non-probability surveys, both at the national and state levels, and for the first three years of the pandemic.

Other attempts to track COVID-19 cases have employed cross-sectional (or limited-period) surveys starting in the early stages of the pandemic. Most of them conclude that only a small fraction of COVID-19 cases were captured by institutional testing, consistent with our findings post-February 2022 –and perhaps also during the early weeks of the pandemic ^5^. For example, a study by Gallup suggested that the number of COVID-19 infections on April 3, 2020, would be 2.5 times more than what the official numbers had suggested back then if more people were to get officially tested. Another survey-based study conducted by Qasmieh et al. fielded between 14 – 16 March 2022, asked 1030 adult residents of New York City about COVID-19 testing and related outcomes from January 2022 onwards^27^. They applied representative survey weights like our methodology to estimate the number of infections in New York City. They estimated that 1.8 million adults (95% CI: 1.6 – 2.1 million) had a COVID-19 infection from 1 January to 16 March 2022, compared to 1.1 million cases that our survey numbers suggest for the same period in New York state.

In another online survey-based (N = 97,707) study led by Rader, Gertz, and Brownstein, researchers estimate that “2.6 million cases [95% CI: 1,874.549 to 3,853,341] were diagnosed by at-home tests and not included in the official case count” over the period March 20 – May 21, 2022. Our surveys’ temporal resolution does not allow for a direct comparison, but when scaling our monthly estimates (March estimate*1/3 + April estimate + May estimate*2/3) we estimate that approximately 6 million infections nationally were not included in reported case counts in the same period.

In another survey-based study, Qasmieh et al. estimated COVID-19 cumulative incidence, during the preceding 14-day period (April 23 – May 8), to be 31 times the official case count: 1.5 million (95% CI 1.3-1.8 million) versus 50 thousand^28^. In comparison, our estimates for April – May 2022 suggest that there were 1.2 million cases in New York state, much closer to Qasmieh et al. estimates. Government-led efforts aimed at centralizing information about individual at-home test results include a National Institutes of Health initiative tasked with developing a self-test reporting standard.

We identify large discrepancies in COVID-19 estimates among all three data sources – CSP. JHU, and WW – before May 2020. Both CSP and WW data show significantly higher estimates of COVID-19 activity than those reported by JHU. While testing was very sparse and inconsistent in this period, our estimates align with other attempts, for example ^21^, that have employed statistical corrections, and multiple complementary data streams, to estimate the total number of COVID-19 infections before April 2020. Specifically, Lu et al. find the cumulative number of suspected (symptomatic, either test-confirmed or not) infections as of April 4, 2020, to be as many as 2.3 to 4.8 million cases, or about 25 times the number of institutionally reported cases in the United States. Our estimates do not point to such high numbers since, by design, our goal was to track only test-confirmed infections.

Since our surveys were not specifically designed to attract the participation of individuals with COVID-19 symptoms or particular interest in COVID-19 more broadly, our infection-rate estimates should be less biased than those obtained from COVID-19 specific surveys such as the “Facebook-CMU” survey or “Outbreaks near me” ^29,30^. Indeed, it has been documented that people who are experiencing symptoms are more motivated to report their experience in surveys ^31,32^ and as such, incidence rates tend to be inflated in disease-specific surveys since fewer healthy individuals participate in them.

It is important to note that one strength of survey-based infection cases surveillance is that it allows for the multivariate collection of disease activity information in parallel to other socio-demographic variables. Institutional data collection, in contrast, rarely allows for access to demographic details of those reported infected and thus precludes looking at subgroup infection rates. Indeed, future studies of the survey data will closely analyze infection trends in different socio-demographic groups.

In future public health crises, survey-based approaches to monitor confirmed infections should be deployed in conjunction with either widely available institutional testing or diagnostic at-home tests. In absence of that, no gold standard will exist to assess the historical validity of survey-based approaches and thus, their robustness and generalizability may be limited.

### Limitations

Our study has multiple limitations. The first one is potential participants’ recall error. We utilized the answers from the most contemporaneous wave to estimate the number of infections in a month to mitigate recall bias. Another potential limitation might be entry errors in low frequency responses. An expected low frequency response in our study was whether the respondent was sick or not at the very beginning of the pandemic^33^. Entry errors for low frequency responses in a (big) ∼20,000-respondent wave could have inflated our test-confirmed infection estimates in the first three months of the pandemic. Although we have a robust statistical power nationally and within big states as shown in the Supplementary Materials, our state-level analyses are far less precise, especially for states where we had smaller sample sizes.

## Conclusion

Our study supports the potential for applying surveys to complement government-led disease surveillance in future public health crises, despite some limitations that may be addressable in future deployments.

## Supporting information

Supplementary_Materials_Santillana_et_al_2024

## Data Availability

Data are available upon reasonable request.

https://www.covidstates.org/

