## Supplementary_Materials_Santillana_et_al_2024 for "Survey data yields improved estimates of test-confirmed COVID-19 cases when rapid at-home tests were massively distributed in the United States"

**Methods**

**Precise wording of questions on survey about COVID-19 infections:**

**Question 1.** Have you been tested for coronavirus (COVID-19)?

- Yes, and I tested positive for COVID-19 at least once
- Yes, and I tested negative for COVID-19 every time
- No, I wanted to but was not able to get a test
- No, I never tried to get tested

If answer is yes for Question 1, then:

**Question 2.** In which months of 2020 were you sick? (Please select all that apply)

- January 2020
- February 2020
- March 2020
- …
- December 2022

**Results**

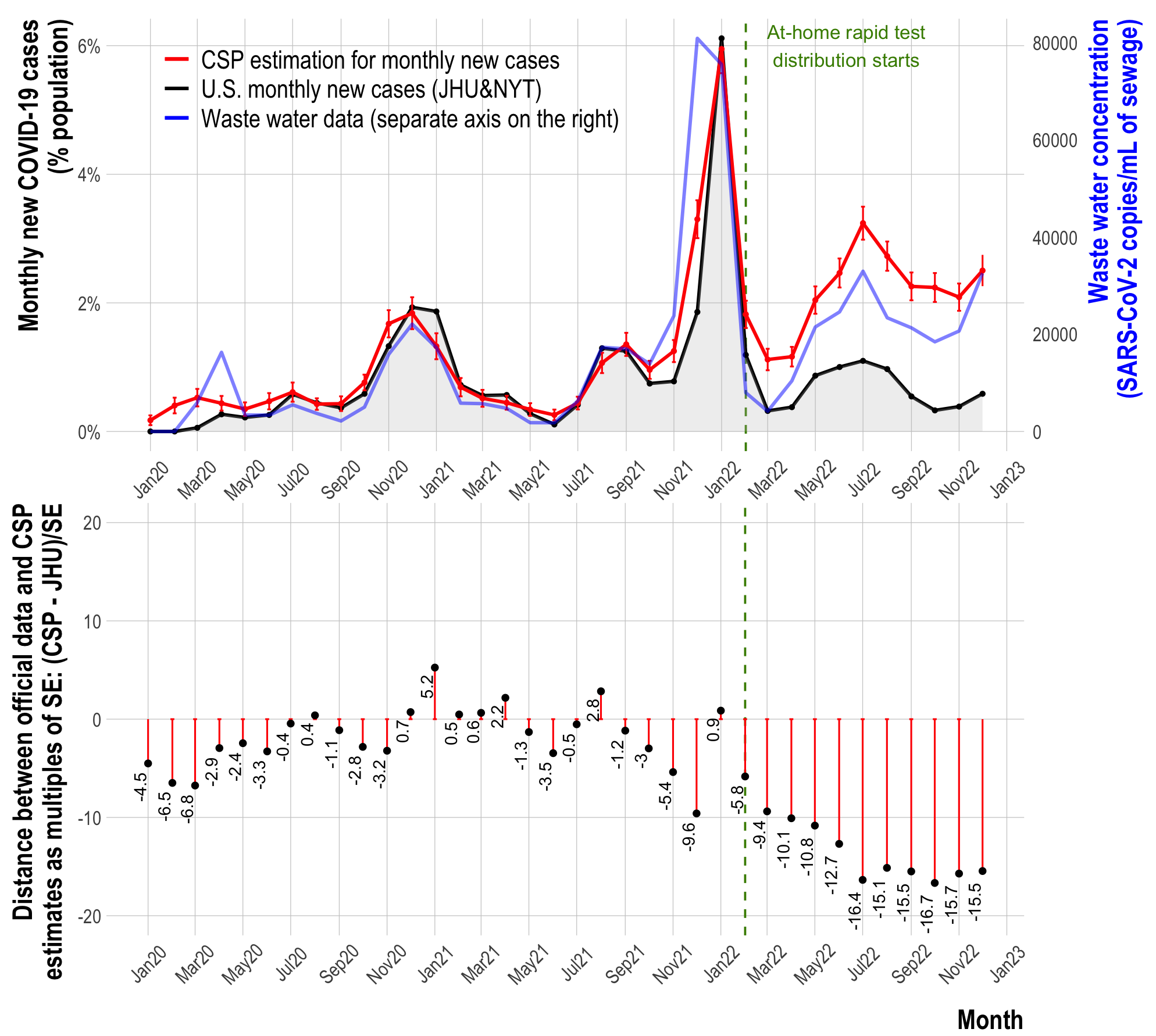

**Figure S1. Top panel:** The percent of respondents in our survey (CSP) who reported having a confirmed COVID-19 infection in each month is shown in red, the institutionally reported percent of individuals infected in each month as monitored by JHU is shown in black, and the wastewater viral concentration of SARS-CoV-2 is shown in blue. A vertical green dashed line shows the time when at-home rapid test were widely delivered in February 2022. **Bottom panel:** differences between CSP and JHU new monthly infections as multiples of the standard error (of means) of the CSP estimates.

| **Period** | **CSP-JHU** | **WW-CSP** | **JHU-WW** |
| --- | --- | --- | --- |
| **Apr 2020 – Jan 2022 (pre-rapid test period)** | 0.882 (SD = 0.073) | 0.614 (SD = 0.374) | 0.652 (SD = 0.290) |
| **Feb 2022 – Jan 2023 (rapid test period)** | 0.48 (SD = 0.227) | 0.470 (SD = 0.314) | 0.146 (SD = 0.322) |

**Table S1.** Average Pearson correlation for all US states between survey test-confirmed infections estimates (CSP), Institutionally reported COVID-19 (JHU), and Wastewater SARS-CoV-2 viral concentrations (WW) in two time periods.

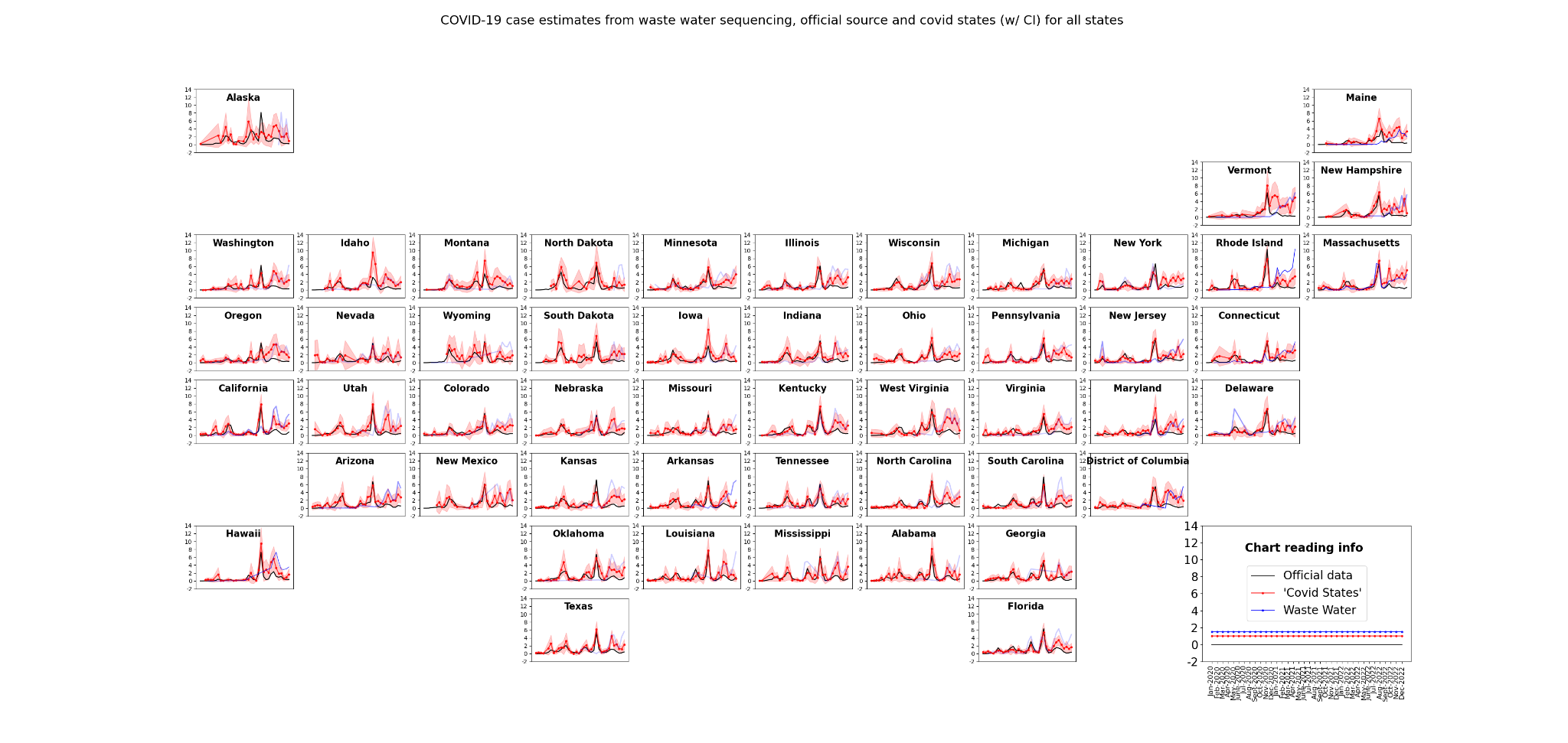

**Figure S2:** COVID-19 case estimates from CSP data (with confidence intervals, in red), concentrations of SARS-CoV-2 in wastewater (blue), and institutional confirmed cases from JHU (black) for all US states. For Waste water concentrations, darker blue colors indicate the presence of higher numbers of observation stations and thus, more robust estimates.

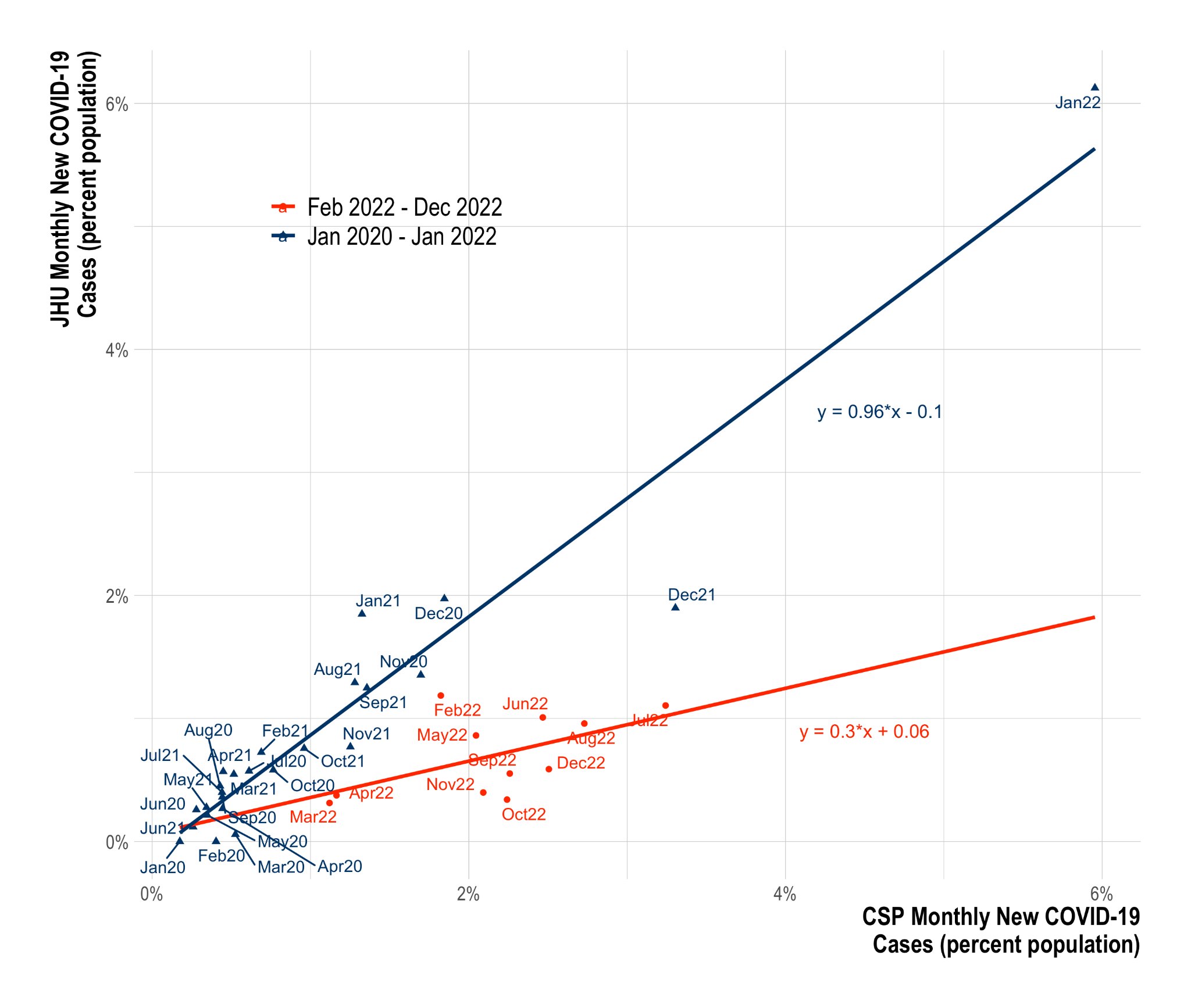

**Figure S3.** Scatter plots of JHU COVID-19 infections vs CSP infections from April 2020 to January 2022 was used for training (blue) and from February 2022 to January 2023 (red). Individually fitted linear regressions for both time periods and their equations are included. Using the regression on the time period from April 2020 to January 2022 as our **interrupted time series** approach, we calculated the infections that would have been observed in the JHU data during the time period February 2022 to January 2023, had rapid at-home tests not been widely distributed and JHU surveillance had not been dramatically reduced.

**State-wise case estimation and data volume**

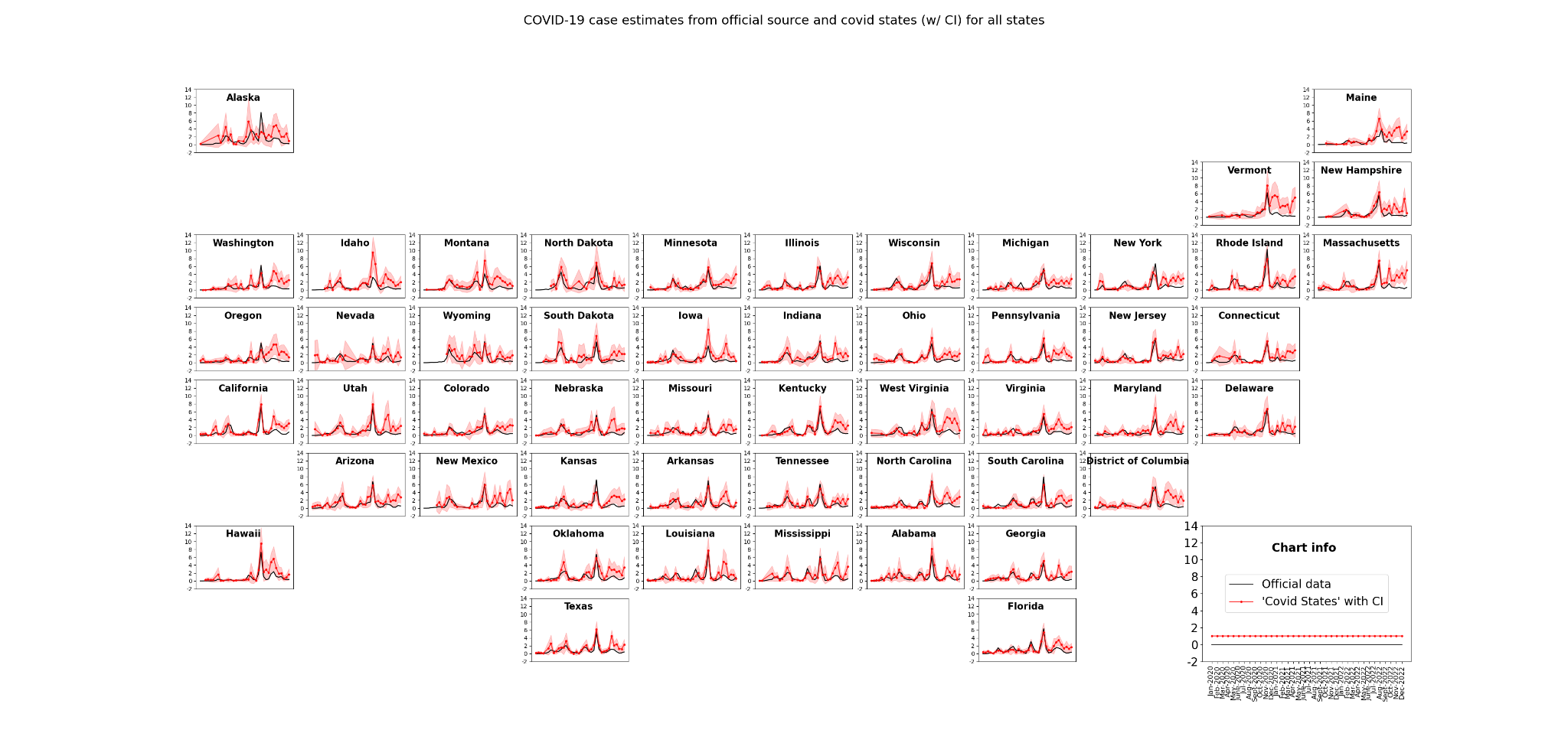

**Figure S4:** COVID-19 case estimates from Covid States Project data (with confidence interval) and official data source for all states

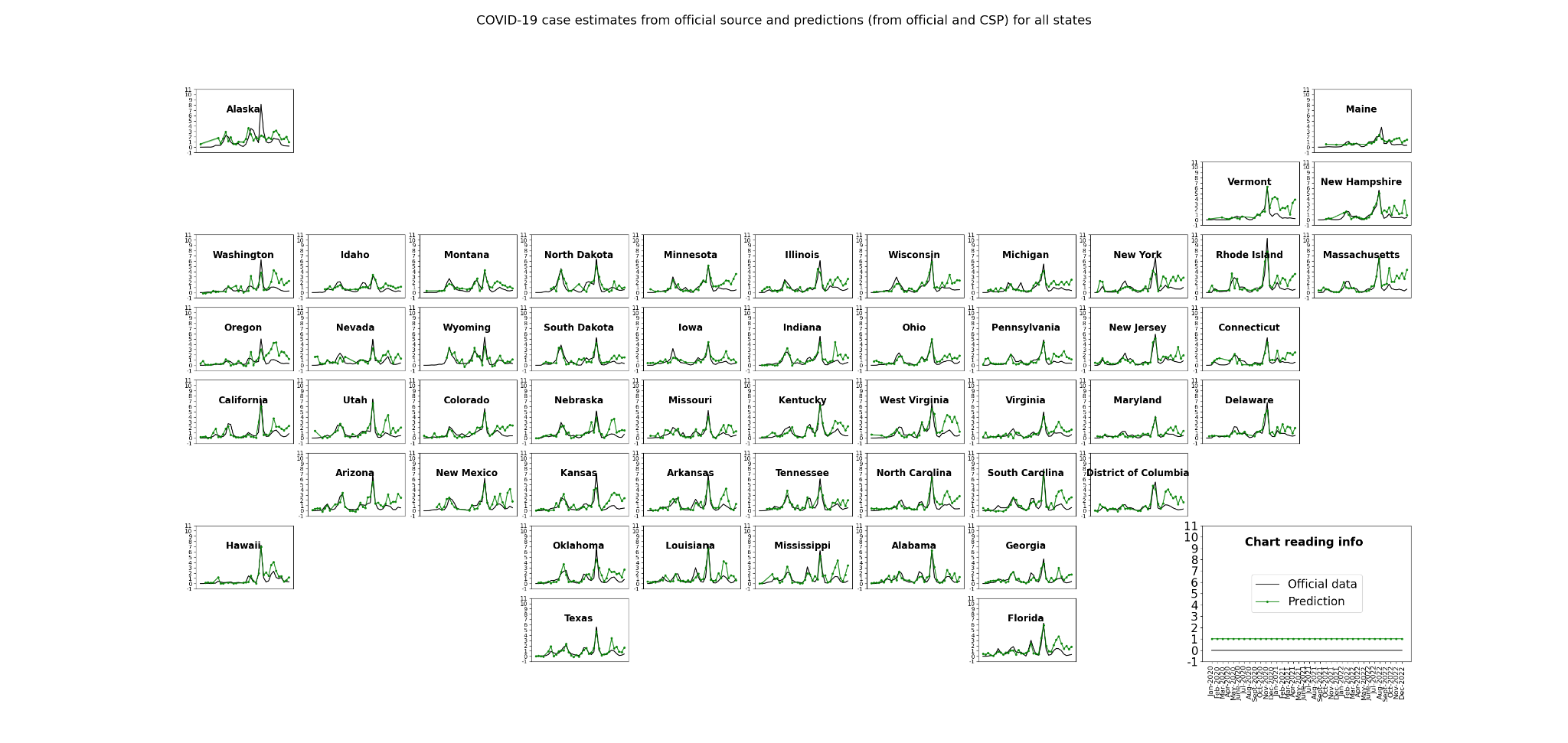

**Figure S5*:*** *Interrupted time-series approach:* COVID-19 case estimates from JHU (black) and predictions using interrupted linear regression relating official source and Covid States Project for all states during April 2020 and January 2022.

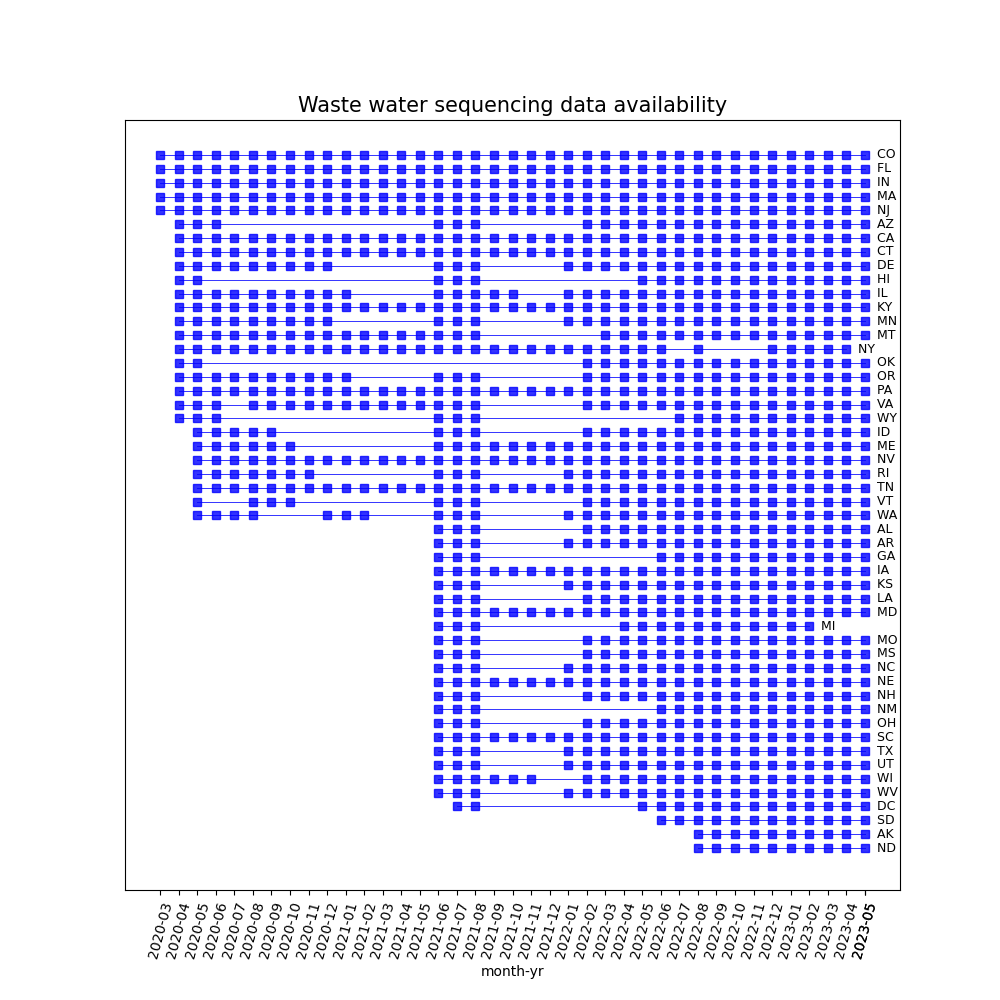

**Figure S6:** Data availability from Biobot’s wastewater sequencing per month in each state. Months with no data has been left blank and those with data has been marked with blue squares.

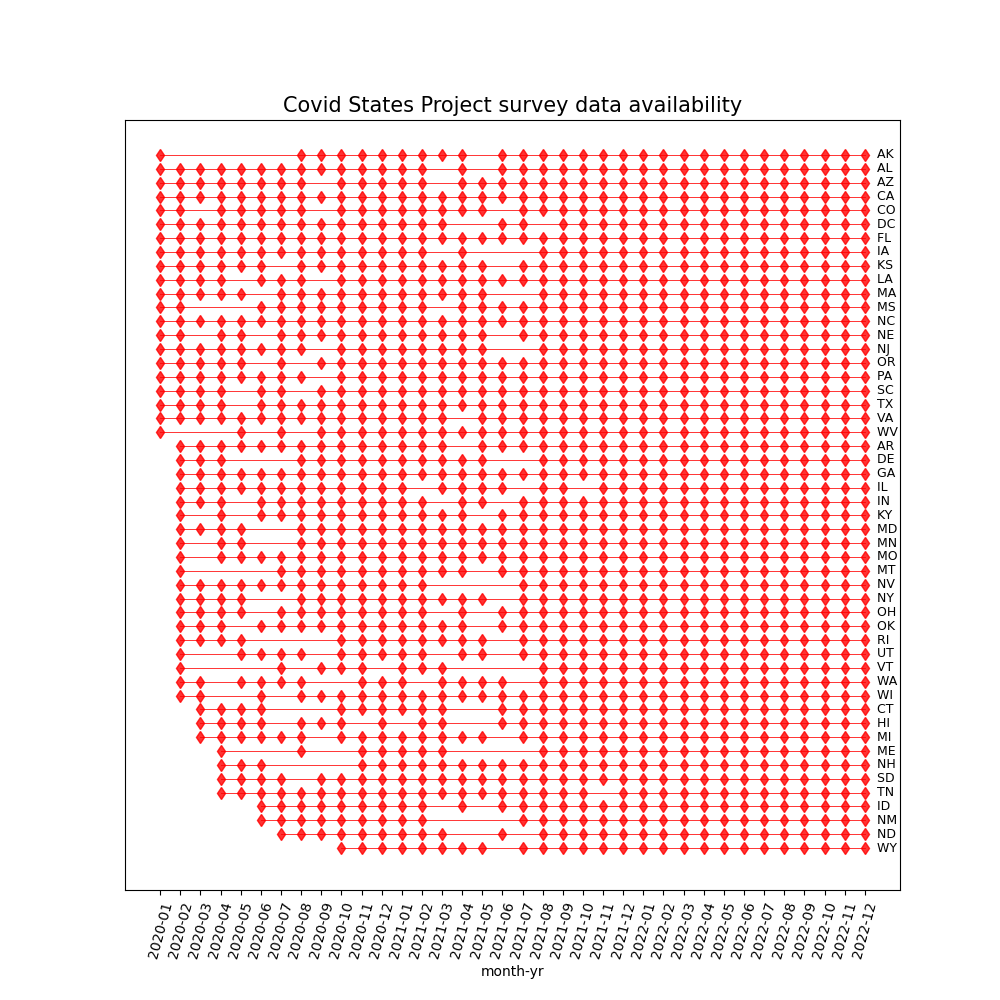

**Figure S7:** Data availability from COVID states project survey per month in each state. Months with no data has been left blank and those with data has been marked with red circles.

**Table S2** State-level pairwise Pearson correlation and p-values between survey test-confirmed infections estimates (CSP), Institutionally reported COVID-19 (JHU), and Wastewater SARS-CoV-2 viral concentrations (WW) in three time periods.

|  | **state** | **CSP - JHU** | **WW - CSP** | **JHU - WW** |
| --- | --- | --- | --- | --- |
| **Apr 2020 – Jan 2023 (full data)** | CO | 0.7236 (p = 2e-06) | 0.7583 (p = 0.0) | 0.3163 (p = 0.072877) |
| **Apr 2020 – Jan 2022 (pre-rapid test period)** | CO | 0.9151 (p = 0.0) | 0.8679 (p = 0.0) | 0.7713 (p = 2.6e-05) |
| **Feb 2022 – Jan 2023 (rapid test period)** | CO | 0.0624 (p = 0.855307) | 0.6247 (p = 0.039877) | -0.0931 (p = 0.785523) |
| **Apr 2020 – Jan 2023 (full data)** | FL | 0.7779 (p = 0.0) | 0.781 (p = 0.0) | 0.4276 (p = 0.013055) |
| **Apr 2020 – Jan 2022 (pre-rapid test period)** | FL | 0.9332 (p = 0.0) | 0.771 (p = 2.7e-05) | 0.741 (p = 8e-05) |
| **Feb 2022 – Jan 2023 (rapid test period)** | FL | 0.8204 (p = 0.001977) | 0.8227 (p = 0.001874) | 0.5378 (p = 0.087921) |
| **Apr 2020 – Jan 2023 (full data)** | IN | 0.7062 (p = 4e-06) | 0.5331 (p = 0.001403) | 0.0528 (p = 0.770285) |
| **Apr 2020 – Jan 2022 (pre-rapid test period)** | IN | 0.9233 (p = 0.0) | 0.7829 (p = 1.7e-05) | 0.811 (p = 5e-06) |
| **Feb 2022 – Jan 2023 (rapid test period)** | IN | 0.473 (p = 0.141719) | 0.6645 (p = 0.025714) | 0.5626 (p = 0.071605) |
| **Apr 2020 – Jan 2023 (full data)** | MA | 0.6321 (p = 7.9e-05) | 0.852 (p = 0.0) | 0.6843 (p = 1.1e-05) |
| **Apr 2020 – Jan 2022 (pre-rapid test period)** | MA | 0.9425 (p = 0.0) | 0.868 (p = 0.0) | 0.8645 (p = 0.0) |
| **Feb 2022 – Jan 2023 (rapid test period)** | MA | 0.3776 (p = 0.252319) | 0.7323 (p = 0.010383) | -0.0626 (p = 0.854949) |
| **Apr 2020 – Jan 2023 (full data)** | NJ | 0.805 (p = 0.0) | 0.3883 (p = 0.025526) | 0.0624 (p = 0.730301) |
| **Apr 2020 – Jan 2022 (pre-rapid test period)** | NJ | 0.9414 (p = 0.0) | 0.2781 (p = 0.21008) | 0.2159 (p = 0.334449) |
| **Feb 2022 – Jan 2023 (rapid test period)** | NJ | 0.0577 (p = 0.8662) | 0.4139 (p = 0.205712) | 0.042 (p = 0.902506) |
| **Apr 2020 – Jan 2023 (full data)** | CA | 0.752 (p = 0.0) | 0.5897 (p = 0.000305) | 0.2327 (p = 0.192554) |
| **Apr 2020 – Jan 2022 (pre-rapid test period)** | CA | 0.865 (p = 0.0) | 0.7254 (p = 0.000133) | 0.7878 (p = 1.3e-05) |
| **Feb 2022 – Jan 2023 (rapid test period)** | CA | 0.414 (p = 0.205536) | 0.8567 (p = 0.000757) | 0.5687 (p = 0.067881) |
| **Apr 2020 – Jan 2023 (full data)** | CT | 0.6849 (p = 1.1e-05) | 0.5595 (p = 0.000712) | 0.0747 (p = 0.679505) |
| **Apr 2020 – Jan 2022 (pre-rapid test period)** | CT | 0.874 (p = 0.0) | 0.5454 (p = 0.008662) | 0.5765 (p = 0.004978) |
| **Feb 2022 – Jan 2023 (rapid test period)** | CT | 0.4207 (p = 0.197546) | 0.522 (p = 0.099571) | -0.0726 (p = 0.83195) |
| **Apr 2020 – Jan 2023 (full data)** | KY | 0.7944 (p = 0.0) | 0.7994 (p = 0.0) | 0.4976 (p = 0.003215) |
| **Apr 2020 – Jan 2022 (pre-rapid test period)** | KY | 0.9165 (p = 0.0) | 0.9133 (p = 0.0) | 0.8949 (p = 0.0) |
| **Feb 2022 – Jan 2023 (rapid test period)** | KY | 0.6155 (p = 0.043819) | 0.7627 (p = 0.006336) | 0.3957 (p = 0.228368) |
| **Apr 2020 – Jan 2023 (full data)** | PA | 0.715 (p = 3e-06) | 0.7822 (p = 0.0) | 0.3884 (p = 0.025489) |
| **Apr 2020 – Jan 2022 (pre-rapid test period)** | PA | 0.954 (p = 0.0) | 0.8659 (p = 0.0) | 0.8317 (p = 2e-06) |
| **Feb 2022 – Jan 2023 (rapid test period)** | PA | 0.502 (p = 0.115618) | 0.4562 (p = 0.158407) | 0.2932 (p = 0.381495) |
| **Apr 2020 – Jan 2023 (full data)** | NV | 0.5962 (p = 0.000251) | 0.8064 (p = 0.0) | 0.7173 (p = 3e-06) |
| **Apr 2020 – Jan 2022 (pre-rapid test period)** | NV | 0.8533 (p = 0.0) | 0.8077 (p = 5e-06) | 0.8529 (p = 0.0) |
| **Feb 2022 – Jan 2023 (rapid test period)** | NV | 0.5533 (p = 0.077471) | 0.8126 (p = 0.002367) | 0.4995 (p = 0.117759) |
| **Apr 2020 – Jan 2023 (full data)** | TN | 0.7889 (p = 0.0) | 0.6269 (p = 9.5e-05) | 0.5667 (p = 0.000585) |
| **Apr 2020 – Jan 2022 (pre-rapid test period)** | TN | 0.8602 (p = 0.0) | 0.7261 (p = 0.00013) | 0.8706 (p = 0.0) |
| **Feb 2022 – Jan 2023 (rapid test period)** | TN | 0.7364 (p = 0.009749) | 0.4238 (p = 0.19395) | 0.2652 (p = 0.430663) |
| **Apr 2020 – Jan 2023 (full data)** | NY | 0.5794 (p = 0.00041) | 0.4821 (p = 0.004498) | 0.6896 (p = 9e-06) |
| **Apr 2020 – Jan 2022 (pre-rapid test period)** | NY | 0.8133 (p = 4e-06) | 0.7915 (p = 1.1e-05) | 0.6958 (p = 0.000323) |
| **Feb 2022 – Jan 2023 (rapid test period)** | NY | 0.4346 (p = 0.181646) | Insufficient data | Insufficient data |
| **Apr 2020 – Jan 2023 (full data)** | IL | 0.6639 (p = 2.5e-05) | 0.4203 (p = 0.014888) | -0.0391 (p = 0.828884) |
| **Apr 2020 – Jan 2022 (pre-rapid test period)** | IL | 0.8307 (p = 2e-06) | -0.0372 (p = 0.869481) | 0.1542 (p = 0.493137) |
| **Feb 2022 – Jan 2023 (rapid test period)** | IL | 0.6073 (p = 0.047527) | 0.5006 (p = 0.116792) | 0.3405 (p = 0.305485) |
| **Apr 2020 – Jan 2023 (full data)** | MT | 0.7966 (p = 0.0) | 0.1044 (p = 0.563299) | 0.0601 (p = 0.739731) |
| **Apr 2020 – Jan 2022 (pre-rapid test period)** | MT | 0.9095 (p = 0.0) | Insufficient data | Insufficient data |
| **Feb 2022 – Jan 2023 (rapid test period)** | MT | 0.6307 (p = 0.037483) | -0.4499 (p = 0.165025) | -0.4599 (p = 0.154633) |
| **Apr 2020 – Jan 2023 (full data)** | VA | 0.679 (p = 1.4e-05) | 0.4774 (p = 0.004958) | -0.0232 (p = 0.897852) |
| **Apr 2020 – Jan 2022 (pre-rapid test period)** | VA | 0.8535 (p = 0.0) | -0.3065 (p = 0.165369) | 0.0564 (p = 0.803179) |
| **Feb 2022 – Jan 2023 (rapid test period)** | VA | 0.7241 (p = 0.011743) | 0.4751 (p = 0.139761) | 0.1832 (p = 0.589743) |
| **Apr 2020 – Jan 2023 (full data)** | ME | 0.5703 (p = 0.000531) | 0.6131 (p = 0.000148) | 0.0276 (p = 0.878888) |
| **Apr 2020 – Jan 2022 (pre-rapid test period)** | ME | Insufficient data | Insufficient data | Insufficient data |
| **Feb 2022 – Jan 2023 (rapid test period)** | ME | 0.2653 (p = 0.430362) | 0.2518 (p = 0.455069) | -0.4202 (p = 0.198149) |
| **Apr 2020 – Jan 2023 (full data)** | DE | 0.7908 (p = 0.0) | 0.1926 (p = 0.282968) | 0.0551 (p = 0.760699) |
| **Apr 2020 – Jan 2022 (pre-rapid test period)** | DE | 0.8957 (p = 0.0) | Insufficient data | Insufficient data |
| **Feb 2022 – Jan 2023 (rapid test period)** | DE | 0.4068 (p = 0.214426) | 0.4697 (p = 0.144952) | 0.3762 (p = 0.254185) |
| **Apr 2020 – Jan 2023 (full data)** | MN | 0.6939 (p = 8e-06) | 0.4326 (p = 0.011913) | -0.1146 (p = 0.525347) |
| **Apr 2020 – Jan 2022 (pre-rapid test period)** | MN | 0.9526 (p = 0.0) | Insufficient data | Insufficient data |
| **Feb 2022 – Jan 2023 (rapid test period)** | MN | 0.2159 (p = 0.523648) | 0.3322 (p = 0.318164) | -0.1257 (p = 0.71262) |
| **Apr 2020 – Jan 2023 (full data)** | OR | 0.4321 (p = 0.012034) | 0.6671 (p = 2.2e-05) | -0.0153 (p = 0.93258) |
| **Apr 2020 – Jan 2022 (pre-rapid test period)** | OR | 0.7893 (p = 1.3e-05) | Insufficient data | Insufficient data |
| **Feb 2022 – Jan 2023 (rapid test period)** | OR | 0.3145 (p = 0.346187) | 0.4303 (p = 0.18647) | 0.0012 (p = 0.997249) |
| **Apr 2020 – Jan 2023 (full data)** | RI | 0.7828 (p = 0.0) | Insufficient data | Insufficient data |
| **Apr 2020 – Jan 2022 (pre-rapid test period)** | RI | 0.9012 (p = 0.0) | Insufficient data | Insufficient data |
| **Feb 2022 – Jan 2023 (rapid test period)** | RI | 0.1323 (p = 0.698122) | 0.8303 (p = 0.001555) | -0.0747 (p = 0.827154) |
| **Apr 2020 – Jan 2023 (full data)** | WA | 0.5164 (p = 0.002096) | Insufficient data | Insufficient data |
| **Apr 2020 – Jan 2022 (pre-rapid test period)** | WA | 0.7744 (p = 2.3e-05) | Insufficient data | Insufficient data |
| **Feb 2022 – Jan 2023 (rapid test period)** | WA | 0.2944 (p = 0.379483) | 0.6755 (p = 0.022531) | -0.1275 (p = 0.708757) |
| **Apr 2020 – Jan 2023 (full data)** | ID | 0.7438 (p = 1e-06) | Insufficient data | Insufficient data |
| **Apr 2020 – Jan 2022 (pre-rapid test period)** | ID | 0.8119 (p = 4e-06) | Insufficient data | Insufficient data |
| **Feb 2022 – Jan 2023 (rapid test period)** | ID | 0.8588 (p = 0.00071) | 0.3319 (p = 0.318764) | -0.0347 (p = 0.919299) |
| **Apr 2020 – Jan 2023 (full data)** | IA | 0.7188 (p = 2e-06) | Insufficient data | Insufficient data |
| **Apr 2020 – Jan 2022 (pre-rapid test period)** | IA | 0.8597 (p = 0.0) | Insufficient data | Insufficient data |
| **Feb 2022 – Jan 2023 (rapid test period)** | IA | 0.6457 (p = 0.031862) | 0.329 (p = 0.323265) | 0.0882 (p = 0.796618) |
| **Apr 2020 – Jan 2023 (full data)** | MD | 0.7618 (p = 0.0) | Insufficient data | Insufficient data |
| **Apr 2020 – Jan 2022 (pre-rapid test period)** | MD | 0.9291 (p = 0.0) | Insufficient data | Insufficient data |
| **Feb 2022 – Jan 2023 (rapid test period)** | MD | 0.3351 (p = 0.313778) | 0.4576 (p = 0.156968) | -0.024 (p = 0.944097) |
| **Apr 2020 – Jan 2023 (full data)** | NE | 0.6265 (p = 9.6e-05) | Insufficient data | Insufficient data |
| **Apr 2020 – Jan 2022 (pre-rapid test period)** | NE | 0.8651 (p = 0.0) | Insufficient data | Insufficient data |
| **Feb 2022 – Jan 2023 (rapid test period)** | NE | 0.3343 (p = 0.315053) | 0.5225 (p = 0.099164) | 0.0034 (p = 0.992116) |
| **Apr 2020 – Jan 2023 (full data)** | SC | 0.7537 (p = 0.0) | Insufficient data | Insufficient data |
| **Apr 2020 – Jan 2022 (pre-rapid test period)** | SC | 0.934 (p = 0.0) | Insufficient data | Insufficient data |
| **Feb 2022 – Jan 2023 (rapid test period)** | SC | 0.4875 (p = 0.128272) | 0.9071 (p = 0.000116) | 0.6123 (p = 0.045226) |
| **Apr 2020 – Jan 2023 (full data)** | VT | 0.6307 (p = 8.3e-05) | Insufficient data | Insufficient data |
| **Apr 2020 – Jan 2022 (pre-rapid test period)** | VT | Insufficient data | Insufficient data | Insufficient data |
| **Feb 2022 – Jan 2023 (rapid test period)** | VT | 0.3524 (p = 0.287882) | -0.2982 (p = 0.373104) | -0.7811 (p = 0.004536) |
| **Apr 2020 – Jan 2023 (full data)** | AZ | 0.7387 (p = 1e-06) | Insufficient data | Insufficient data |
| **Apr 2020 – Jan 2022 (pre-rapid test period)** | AZ | 0.9103 (p = 0.0) | Insufficient data | Insufficient data |
| **Feb 2022 – Jan 2023 (rapid test period)** | AZ | 0.1653 (p = 0.627195) | 0.8001 (p = 0.003103) | 0.1438 (p = 0.673149) |
| **Apr 2020 – Jan 2023 (full data)** | WI | 0.7765 (p = 0.0) | Insufficient data | Insufficient data |
| **Apr 2020 – Jan 2022 (pre-rapid test period)** | WI | 0.9215 (p = 0.0) | Insufficient data | Insufficient data |
| **Feb 2022 – Jan 2023 (rapid test period)** | WI | 0.2774 (p = 0.408833) | 0.8324 (p = 0.001475) | 0.1852 (p = 0.585717) |
| **Apr 2020 – Jan 2023 (full data)** | AR | 0.7431 (p = 1e-06) | Insufficient data | Insufficient data |
| **Apr 2020 – Jan 2022 (pre-rapid test period)** | AR | 0.8834 (p = 0.0) | Insufficient data | Insufficient data |
| **Feb 2022 – Jan 2023 (rapid test period)** | AR | 0.758 (p = 0.006871) | -0.0763 (p = 0.823611) | -0.1209 (p = 0.723271) |
| **Apr 2020 – Jan 2023 (full data)** | KS | 0.5675 (p = 0.000573) | Insufficient data | Insufficient data |
| **Apr 2020 – Jan 2022 (pre-rapid test period)** | KS | 0.7957 (p = 1e-05) | Insufficient data | Insufficient data |
| **Feb 2022 – Jan 2023 (rapid test period)** | KS | 0.3634 (p = 0.271923) | 0.7607 (p = 0.006563) | 0.4189 (p = 0.19967) |
| **Apr 2020 – Jan 2023 (full data)** | NC | 0.7245 (p = 2e-06) | Insufficient data | Insufficient data |
| **Apr 2020 – Jan 2022 (pre-rapid test period)** | NC | 0.9219 (p = 0.0) | Insufficient data | Insufficient data |
| **Feb 2022 – Jan 2023 (rapid test period)** | NC | 0.463 (p = 0.151521) | 0.7768 (p = 0.00492) | 0.353 (p = 0.286984) |
| **Apr 2020 – Jan 2023 (full data)** | TX | 0.8047 (p = 0.0) | Insufficient data | Insufficient data |
| **Apr 2020 – Jan 2022 (pre-rapid test period)** | TX | 0.9207 (p = 0.0) | Insufficient data | Insufficient data |
| **Feb 2022 – Jan 2023 (rapid test period)** | TX | 0.6588 (p = 0.0275) | 0.6221 (p = 0.040975) | 0.2459 (p = 0.466065) |
| **Apr 2020 – Jan 2023 (full data)** | UT | 0.7591 (p = 0.0) | Insufficient data | Insufficient data |
| **Apr 2020 – Jan 2022 (pre-rapid test period)** | UT | 0.9447 (p = 0.0) | Insufficient data | Insufficient data |
| **Feb 2022 – Jan 2023 (rapid test period)** | UT | 0.7162 (p = 0.01316) | 0.5196 (p = 0.10138) | 0.6158 (p = 0.04368) |
| **Apr 2020 – Jan 2023 (full data)** | WV | 0.6368 (p = 6.8e-05) | Insufficient data | Insufficient data |
| **Apr 2020 – Jan 2022 (pre-rapid test period)** | WV | 0.8918 (p = 0.0) | Insufficient data | Insufficient data |
| **Feb 2022 – Jan 2023 (rapid test period)** | WV | 0.6785 (p = 0.021709) | 0.57 (p = 0.067144) | 0.2239 (p = 0.508015) |
| **Apr 2020 – Jan 2023 (full data)** | AL | 0.8425 (p = 0.0) | Insufficient data | Insufficient data |
| **Apr 2020 – Jan 2022 (pre-rapid test period)** | AL | 0.9297 (p = 0.0) | Insufficient data | Insufficient data |
| **Feb 2022 – Jan 2023 (rapid test period)** | AL | 0.7204 (p = 0.012402) | 0.092 (p = 0.787884) | 0.1043 (p = 0.760276) |
| **Apr 2020 – Jan 2023 (full data)** | LA | 0.7408 (p = 1e-06) | Insufficient data | Insufficient data |
| **Apr 2020 – Jan 2022 (pre-rapid test period)** | LA | 0.8853 (p = 0.0) | Insufficient data | Insufficient data |
| **Feb 2022 – Jan 2023 (rapid test period)** | LA | 0.5568 (p = 0.075228) | 0.2716 (p = 0.419066) | 0.4731 (p = 0.14164) |
| **Apr 2020 – Jan 2023 (full data)** | MS | 0.6914 (p = 8e-06) | Insufficient data | Insufficient data |
| **Apr 2020 – Jan 2022 (pre-rapid test period)** | MS | 0.8956 (p = 0.0) | Insufficient data | Insufficient data |
| **Feb 2022 – Jan 2023 (rapid test period)** | MS | 0.4599 (p = 0.15463) | 0.7206 (p = 0.012353) | 0.3361 (p = 0.312168) |
| **Apr 2020 – Jan 2023 (full data)** | MO | 0.6598 (p = 3e-05) | Insufficient data | Insufficient data |
| **Apr 2020 – Jan 2022 (pre-rapid test period)** | MO | 0.8221 (p = 3e-06) | Insufficient data | Insufficient data |
| **Feb 2022 – Jan 2023 (rapid test period)** | MO | 0.4917 (p = 0.124541) | 0.428 (p = 0.189073) | 0.1276 (p = 0.708419) |
| **Apr 2020 – Jan 2023 (full data)** | NH | 0.7256 (p = 2e-06) | Insufficient data | Insufficient data |
| **Apr 2020 – Jan 2022 (pre-rapid test period)** | NH | 0.9665 (p = 0.0) | Insufficient data | Insufficient data |
| **Feb 2022 – Jan 2023 (rapid test period)** | NH | -0.1571 (p = 0.644551) | -0.2385 (p = 0.479942) | -0.3909 (p = 0.234609) |
| **Apr 2020 – Jan 2023 (full data)** | OH | 0.8445 (p = 0.0) | Insufficient data | Insufficient data |
| **Apr 2020 – Jan 2022 (pre-rapid test period)** | OH | 0.9667 (p = 0.0) | Insufficient data | Insufficient data |
| **Feb 2022 – Jan 2023 (rapid test period)** | OH | 0.7766 (p = 0.004939) | 0.6322 (p = 0.036889) | 0.4867 (p = 0.128978) |
| **Apr 2020 – Jan 2023 (full data)** | HI | 0.9004 (p = 0.0) | Insufficient data | Insufficient data |
| **Apr 2020 – Jan 2022 (pre-rapid test period)** | HI | 0.9731 (p = 0.0) | Insufficient data | Insufficient data |
| **Feb 2022 – Jan 2023 (rapid test period)** | HI | 0.8599 (p = 0.000687) | 0.348 (p = 0.294342) | 0.4845 (p = 0.131016) |
| **Apr 2020 – Jan 2023 (full data)** | OK | 0.6894 (p = 9e-06) | Insufficient data | Insufficient data |
| **Apr 2020 – Jan 2022 (pre-rapid test period)** | OK | 0.8422 (p = 1e-06) | Insufficient data | Insufficient data |
| **Feb 2022 – Jan 2023 (rapid test period)** | OK | 0.7484 (p = 0.008058) | 0.3519 (p = 0.288587) | 0.5257 (p = 0.096757) |
| **Apr 2020 – Jan 2023 (full data)** | WY | 0.827 (p = 0.0) | Insufficient data | Insufficient data |
| **Apr 2020 – Jan 2022 (pre-rapid test period)** | WY | Insufficient data | Insufficient data | Insufficient data |
| **Feb 2022 – Jan 2023 (rapid test period)** | WY | 0.5097 (p = 0.109263) | Insufficient data | Insufficient data |
| **Apr 2020 – Jan 2023 (full data)** | DC | 0.6422 (p = 5.6e-05) | Insufficient data | Insufficient data |
| **Apr 2020 – Jan 2022 (pre-rapid test period)** | DC | 0.9274 (p = 0.0) | Insufficient data | Insufficient data |
| **Feb 2022 – Jan 2023 (rapid test period)** | DC | 0.6751 (p = 0.022638) | 0.3331 (p = 0.316836) | -0.1383 (p = 0.685046) |
| **Apr 2020 – Jan 2023 (full data)** | GA | 0.7048 (p = 5e-06) | Insufficient data | Insufficient data |
| **Apr 2020 – Jan 2022 (pre-rapid test period)** | GA | 0.8661 (p = 0.0) | Insufficient data | Insufficient data |
| **Feb 2022 – Jan 2023 (rapid test period)** | GA | 0.3228 (p = 0.332991) | Insufficient data | Insufficient data |
| **Apr 2020 – Jan 2023 (full data)** | NM | 0.6437 (p = 5.3e-05) | Insufficient data | Insufficient data |
| **Apr 2020 – Jan 2022 (pre-rapid test period)** | NM | 0.8884 (p = 0.0) | Insufficient data | Insufficient data |
| **Feb 2022 – Jan 2023 (rapid test period)** | NM | 0.2396 (p = 0.477896) | Insufficient data | Insufficient data |
| **Apr 2020 – Jan 2023 (full data)** | MI | 0.6783 (p = 1.4e-05) | Insufficient data | Insufficient data |
| **Apr 2020 – Jan 2022 (pre-rapid test period)** | MI | 0.85 (p = 1e-06) | Insufficient data | Insufficient data |
| **Feb 2022 – Jan 2023 (rapid test period)** | MI | 0.2704 (p = 0.421342) | 0.0894 (p = 0.793821) | -0.1524 (p = 0.654552) |
| **Apr 2020 – Jan 2023 (full data)** | SD | 0.7891 (p = 0.0) | Insufficient data | Insufficient data |
| **Apr 2020 – Jan 2022 (pre-rapid test period)** | SD | 0.8654 (p = 0.0) | Insufficient data | Insufficient data |
| **Feb 2022 – Jan 2023 (rapid test period)** | SD | 0.6975 (p = 0.017039) | Insufficient data | Insufficient data |
| **Apr 2020 – Jan 2023 (full data)** | AK | 0.4565 (p = 0.007577) | Insufficient data | Insufficient data |
| **Apr 2020 – Jan 2022 (pre-rapid test period)** | AK | 0.517 (p = 0.013735) | Insufficient data | Insufficient data |
| **Feb 2022 – Jan 2023 (rapid test period)** | AK | 0.5216 (p = 0.099878) | Insufficient data | Insufficient data |
| **Apr 2020 – Jan 2023 (full data)** | ND | 0.8262 (p = 0.0) | Insufficient data | Insufficient data |
| **Apr 2020 – Jan 2022 (pre-rapid test period)** | ND | 0.8735 (p = 0.0) | Insufficient data | Insufficient data |
| **Feb 2022 – Jan 2023 (rapid test period)** | ND | 0.6444 (p = 0.032354) | Insufficient data | Insufficient data |

**Table S3 Monthly observed COVID-19 cases estimated from survey data for the multiple survey deployments.**

| **Month** | **Corresponding contemporaneous wave** | **Fielding date** | **Sample**  **Size** | **CSP Confirmed COVID cases estimate (% population (SE))** | **Conf int (±)** | **Official new cases (% population)** | **Monthly observed wastewater virus copies (SARS-CoV-2 copies / mL of sewage)** |
| --- | --- | --- | --- | --- | --- | --- | --- |
| 1/1/20 | W5 | 06/12/20 - 06/28/20 | 22905 | 0.17 (0.04) | 0.08 | 0 | 0 |
| 2/1/20 | W5 | 06/12/20 - 06/28/20 | 22905 | 0.4 (0.06) | 0.12 | 0 | 4.351 |
| 3/1/20 | W5 | 06/12/20 - 06/28/20 | 22905 | 0.53 (0.07) | 0.14 | 0.057 | 5964.138 |
| 4/1/20 | W5 | 06/12/20 - 06/28/20 | 22905 | 0.44 (0.06) | 0.11 | 0.267 | 16286.723 |
| 5/1/20 | W5 | 06/12/20 - 06/28/20 | 22905 | 0.35 (0.05) | 0.1 | 0.218 | 3403.032 |
| 6/1/20 | W9 | 08/07/20 - 08/26/20 | 21496 | 0.47 (0.06) | 0.13 | 0.257 | 3350.295 |
| 7/1/20 | W9 | 08/07/20 - 08/26/20 | 21496 | 0.61 (0.08) | 0.15 | 0.578 | 5485.554 |
| 8/1/20 | W10 | 09/04/20 - 09/30/20 | 23050 | 0.43 (0.05) | 0.09 | 0.444 | 3713.291 |
| 9/1/20 | W11 | 10/02/20 - 10/23/20 | 19570 | 0.43 (0.06) | 0.11 | 0.367 | 2158.504 |
| 10/1/20 | W13 | 11/03/20 - 11/30/20 | 26642 | 0.76 (0.06) | 0.12 | 0.586 | 5015.679 |
| 11/1/20 | W14 | 12/16/20 - 01/10/21 | 26113 | 1.67 (0.11) | 0.21 | 1.327 | 15875.933 |
| 12/1/20 | W16 | 02/05/21 - 02/28/21 | 23348 | 1.84 (0.13) | 0.25 | 1.931 | 22093.742 |
| 1/1/21 | W16 | 02/05/21 - 02/28/21 | 23348 | 1.33 (0.10) | 0.2 | 1.867 | 17244.19 |
| 2/1/21 | W17 | 04/01/21 - 05/03/21 | 23718 | 0.69 (0.07) | 0.14 | 0.724 | 5824.943 |
| 3/1/21 | W17 | 04/01/21 - 05/03/21 | 23718 | 0.52 (0.07) | 0.13 | 0.559 | 5721.44 |
| 4/1/21 | W18 | 06/09/21 - 07/15/21 | 22275 | 0.45 (0.06) | 0.11 | 0.567 | 4763.132 |
| 5/1/21 | W18 | 06/09/21 - 07/15/21 | 22275 | 0.34 (0.05) | 0.1 | 0.276 | 1804.246 |
| 6/1/21 | W19 | 08/26/21 - 09/27/21 | 23938 | 0.26 (0.04) | 0.08 | 0.109 | 1792.336 |
| 7/1/21 | W19 | 08/26/21 - 09/27/21 | 23938 | 0.44 (0.05) | 0.1 | 0.415 | 6299.917 |
| 8/1/21 | W19 | 08/26/21 - 09/27/21 | 23938 | 1.07 (0.08) | 0.16 | 1.295 | 17335.669 |
| 9/1/21 | W20 | 11/03/21 - 12/02/21 | 24623 | 1.35 (0.09) | 0.18 | 1.247 | 16999.981 |
| 10/1/21 | W20 | 11/03/21 - 12/02/21 | 24623 | 0.96 (0.07) | 0.14 | 0.746 | 13916.669 |
| 11/1/21 | W21 | 12/22/21 - 01/24/22 | 25358 | 1.25 (0.09) | 0.17 | 0.779 | 23814.523 |
| 12/1/21 | W22 | 03/02/22 - 04/09/22 | 23376 | 3.3 (0.15) | 0.29 | 1.858 | 80914.958 |
| 1/1/22 | W22 | 03/02/22 - 04/09/22 | 23376 | 5.95 (0.19) | 0.38 | 6.114 | 75543.053 |
| 2/1/22 | W22 | 03/02/22 - 04/09/22 | 23376 | 1.82 (0.11) | 0.21 | 1.191 | 7985.319 |
| 3/1/22 | W23 | 06/08/22 - 07/05/22 | 24625 | 1.12 (0.09) | 0.17 | 0.32 | 4155.093 |
| 4/1/22 | W23 | 06/08/22 - 07/05/22 | 24625 | 1.16 (0.08) | 0.15 | 0.378 | 10418.822 |
| 5/1/22 | W23 | 06/08/22 - 07/05/22 | 24625 | 2.04 (0.11) | 0.21 | 0.868 | 21505.548 |
| 6/1/22 | W24 | 08/11/22 - 09/11/22 | 26557 | 2.46 (0.12) | 0.23 | 1.002 | 24605.985 |
| 7/1/22 | W24 | 08/11/22 - 09/11/22 | 26557 | 3.24 (0.13) | 0.26 | 1.098 | 32976.992 |
| 8/1/22 | W25 | 10/06/22 - 11/09/22 | 25964 | 2.73 (0.12) | 0.23 | 0.97 | 23397.223 |
| 9/1/22 | W25 | 10/06/22 - 11/09/22 | 25964 | 2.26 (0.11) | 0.22 | 0.545 | 21324.956 |
| 10/1/22 | W26 | 12/22/22 - 01/17/23 | 24957 | 2.24 (0.11) | 0.22 | 0.328 | 18449.675 |
| 11/1/22 | W26 | 12/22/22 - 01/17/23 | 24957 | 2.09 (0.11) | 0.21 | 0.386 | 20673.142 |
| 12/1/22 | W26 | 12/22/22 - 01/17/23 | 24957 | 2.5 (0.12) | 0.24 | 0.586 | 32559.616 |

**Table S4:** The differences in number of cases recorded during the period after rapid tests were deployed on ground (Feb’22 to Dec’22) between Official data source (New York Times) and Covid States survey and prediction obtained by training a linear regression using Covid states.

|  | **state** | **CSP to Official difference in cases** | **Prediction to Official difference in cases** |
| --- | --- | --- | --- |
| **0** | AL | 526990.2407286850 | 339167.7661272380 |
| **1** | AK | 130722.77686534600 | 70925.83735815010 |
| **2** | AZ | 1163460.812393650 | 939389.6976879230 |
| **3** | AR | 331598.5153564970 | 310298.39594262800 |
| **4** | CA | 6333548.762619290 | 3956958.538897410 |
| **5** | CO | 709087.9785993950 | 649805.9267643760 |
| **6** | CT | 582022.3111240730 | 421152.6409606460 |
| **7** | DE | 114836.66901829700 | 94628.34574050710 |
| **8** | DC | 146331.68562350000 | 123482.81987125900 |
| **9** | FL | 2649599.595303260 | 3024054.5740217500 |
| **10** | GA | 1281834.23527784 | 847755.7283917550 |
| **11** | HI | 254350.86896302200 | 143971.60473489300 |
| **12** | ID | 373036.4852108260 | 133555.09081505300 |
| **13** | IL | 2160972.25154087 | 1596224.6016380500 |
| **14** | IN | 1046717.0746397500 | 819152.6839750710 |
| **15** | IA | 498271.9393265960 | 283134.3278201050 |
| **16** | KS | 435905.8917539130 | 469177.11598783900 |
| **17** | KY | 669161.2321769830 | 527628.9195764120 |
| **18** | LA | 528896.3040216120 | 467074.23265232600 |
| **19** | ME | 330705.8162441610 | 70882.51071924790 |
| **20** | MD | 990107.8256367590 | 437804.18152309500 |
| **21** | MA | 1879799.094044760 | 1556633.1033855300 |
| **22** | MI | 1494072.052463090 | 1234583.1526418400 |
| **23** | MN | 987538.7609220750 | 852918.7782339500 |
| **24** | MS | 390256.87324185500 | 349246.0755054090 |
| **25** | MO | 661258.7671207210 | 499102.54315186600 |
| **26** | MT | 185205.11935427600 | 95067.28512833800 |
| **27** | NE | 275152.0241479130 | 215930.52934384500 |
| **28** | NV | 394434.1017132090 | 307211.98362026000 |
| **29** | NH | 240680.32445473500 | 182951.9719613760 |
| **30** | NJ | 1126418.2797025100 | 916286.2946430220 |
| **31** | NM | 362858.2912353530 | 293250.7174076290 |
| **32** | NY | 3370318.1461734700 | 3113796.2989901000 |
| **33** | NC | 1743489.3303337700 | 1705378.2399059200 |
| **34** | ND | 81468.70089653900 | 42395.14151424800 |
| **35** | OH | 1580612.9193829400 | 1101912.833284720 |
| **36** | OK | 773118.397172827 | 557320.7748654880 |
| **37** | OR | 970239.3458168210 | 855289.4586586090 |
| **38** | PA | 2273632.831496540 | 1519336.359492450 |
| **39** | RI | 156100.9546575740 | 173908.50290833200 |
| **40** | SC | 611823.517653686 | 792150.9439946130 |
| **41** | SD | 133829.881385874 | 77775.3258424032 |
| **42** | TN | 717906.5104187630 | 552970.7700769660 |
| **43** | TX | 3700810.7438716500 | 2104511.527014770 |
| **44** | UT | 610019.1389372450 | 473928.36431214400 |
| **45** | VT | 220597.6522698980 | 163321.01305611200 |
| **46** | VA | 1474063.6320805200 | 932795.0678966490 |
| **47** | WA | 1440193.745535710 | 1159384.3790950000 |
| **48** | WV | 405224.1315189560 | 375163.860850622 |
| **49** | WI | 902308.1077957830 | 719750.8848672150 |
| **50** | WY | 59558.02379955020 | 16852.325126042100 |

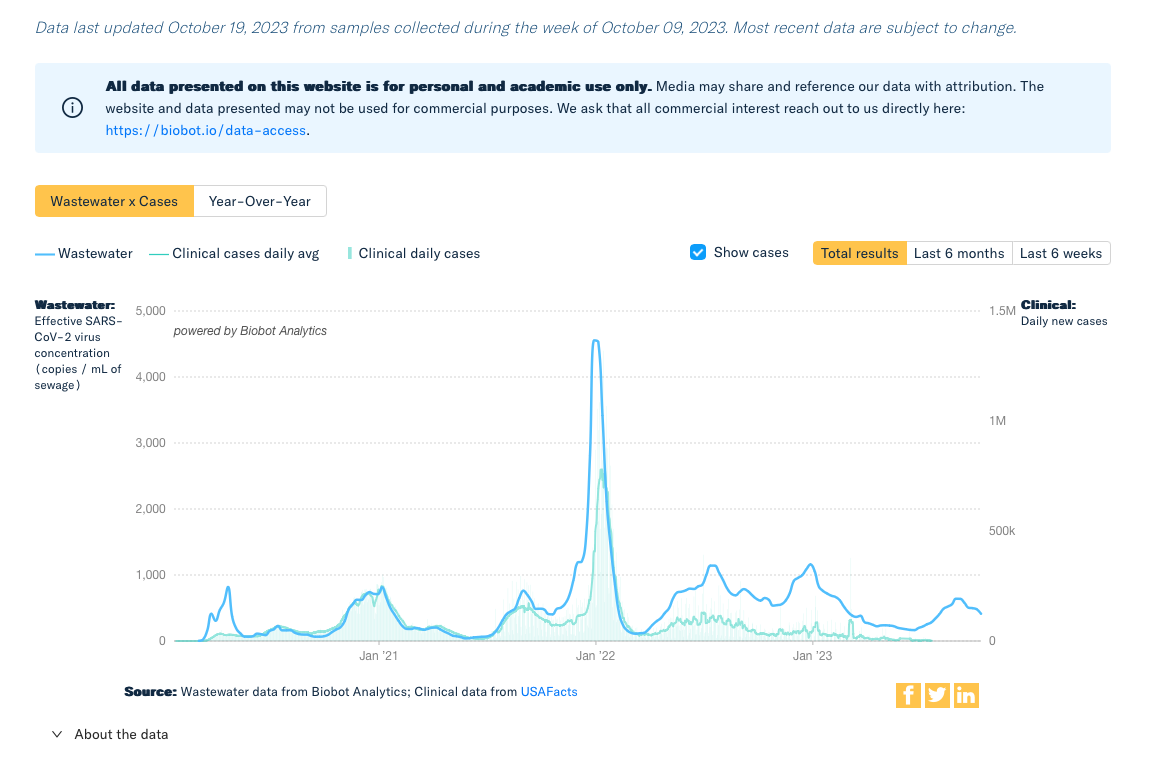

**Figure S8:** Case estimation performed by BioBot using viral concentration in wastewater.

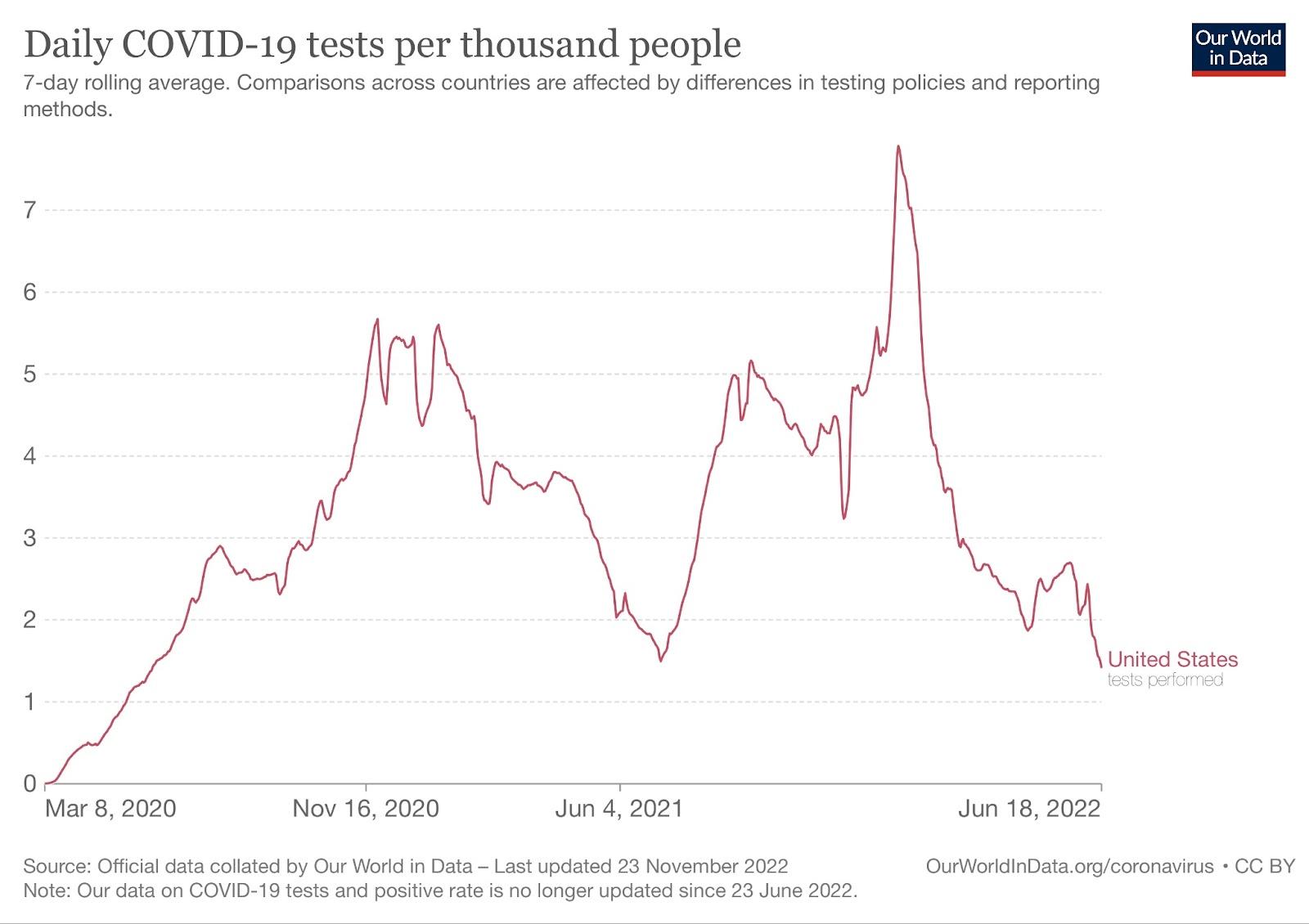

**Figure S9.** Daily COVID-19 tests administered in the United States per thousand people. The data and the figure are from Our World In Data.

**Infection Curves Sensitivity Analysis.**

In order to assess the extent to which our results would vary when including repeat respondents (and potential repeat infections in our analysis), we conducted multiple experiments where we estimated the number of infections in a given point in time, by only including repeat respondents only once (chosen randomly) in our longitudinal analysis. The following plots show that the resulting infections curves estimated from these experiments are very similar to the one that was obtained from including repeat respondents.

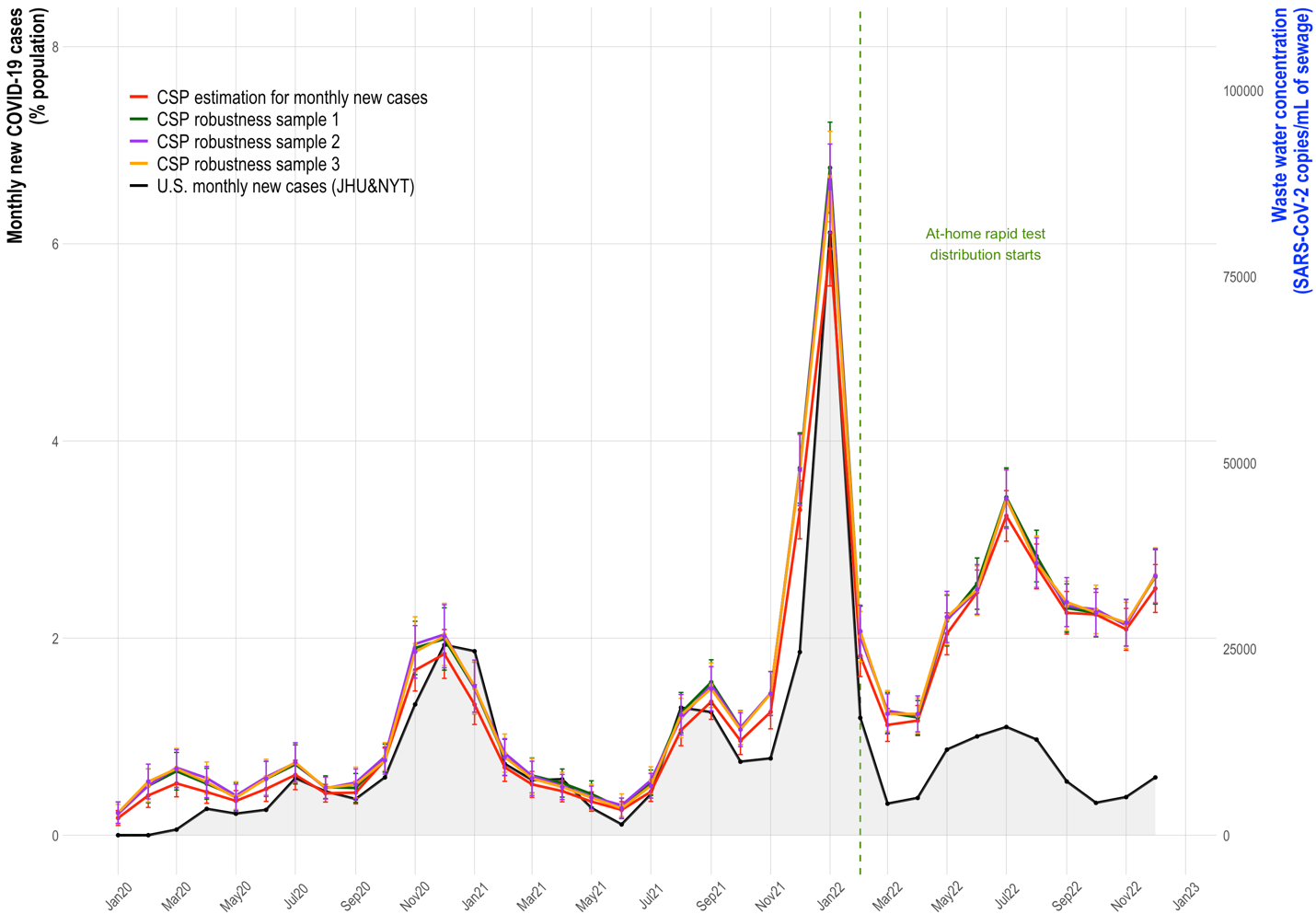

**Figure S10.** Sensitivity analysis of infection curves obtained by only including repeat respondents only once (chosen randomly) in our longitudinal analysis.
